# SVRare: discovering disease-causing structural variants in the 100K Genomes Project

**DOI:** 10.1101/2021.10.15.21265069

**Authors:** Jing Yu, Anita Szabo, Alistair T Pagnamenta, Ahmed Shalaby, Edoardo Giacopuzzi, Jenny Taylor, Deborah Shears, Nikolas Pontikos, Genevieve Wright, Michel Michaelides, Stephanie Halford, Susan Downes, Genomics England Research Consortium

**Author notes:** Corresponding author: Dr Jing Yu.

## Abstract

Discovery of disease-causing structural variants (dcSV) from whole genome sequencing data is difficult due to high number of false positives and a lack of efficient way to estimate allele frequency. Here we introduce SVRare, an application that aggregates structural variants (SV) called by other tools, and efficiently annotates rare SVs to aid dcSVs discovery.

Applied in the Genomics England (GEL) research environment to data from the 100K Genomes Project, SVRare aggregated 554,060,126 SVs called by Manta and Canvas in all the 71,408 participants in the rare-disease arm. From a pilot study of 4313 families, SVRare identified 36 novel protein-coding disrupting SVs on diagnostic grade genes that may explain proband’s phenotype. It is estimated that SVRare can increase SV-based diagnosis yield by at least 4-fold. We also performed a genome-wide association study, and uncovered clusters of dcSVs in genes with known pathogenicity, such as *PKD1/2* - cystic kidney diseases and *LDLR* - familial hypercholesterolaemia.

## Introduction

Structural variants (SVs) are typically defined as DNA changes that extend to at least 50 nucleotides. A wide range of different types of SVs have been shown to cause rare inherited diseases, such as deletion, duplication, inversion, insertion and translocation. Complex SVs involving at least three breakpoint junctions have also been found to cause disease^1^. However, the full contribution of dcSVs to rare diseases is difficult to ascertain, primarily because of a lack of utilities to efficiently prioritise SVs that are potentially disease-causing. This issue is exacerbated for large cohorts of whole genome sequencing (WGS) data.

For the 100K Genome Project (100KGP), a UK wide genome sequencing initiative run by Genomics England (GEL, *The National Genomics Research and Healthcare Knowledgebase v5, Genomics England. doi:10*.*6084/m9*.*figshare*.*4530893*.*v5. 2019*), there were 71,408 participants from 33,924 families in the rare disease arm (as of 20th of June, 2021). The diagnosis pipeline applied by GEL is as shown in Figure 1, where each single nucleotide variant or short indel variant goes through a series of decision stages to be assigned a tier. An essential component of the process is PanelApp, which is a crowd-sourcing knowledgebase that allows virtual gene panels related to human disorders to be created, stored and queried. Each panel may have three categories of genes and regions, and they are coloured according to the evidence available to support aetiology: **green** genes are of diagnostic-grade with high level of evidence to support gene-disease association; **amber** genes have moderate evidence and should not yet be used for genome interpretation; **red** genes have not enough evidence and should not be used for genome interpretation. In GEL, Rare variants of Tier 1 and 2 (protein altering variant in a green gene, on a gene panel from PanelApp applied to the participant) will be further assessed by the NHS Genomics Medical Centres (GMCs) before reporting.

**Figure 1:**
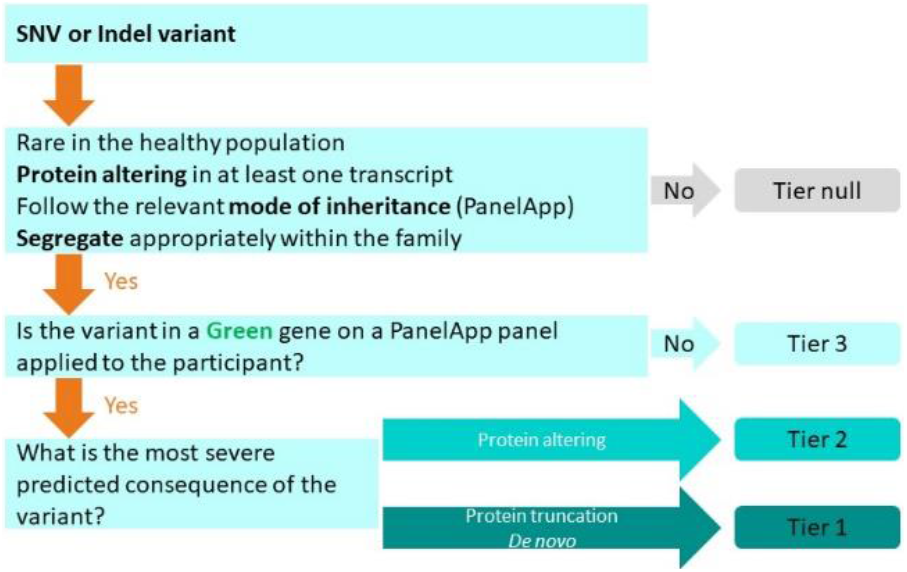
Overview of the GEL”s tiering process for SNV or indel variant. A recreation of the tiering process flowchart from Genomics England “Rare Disease Results Guide”.

GEL also features a copy number variant (CNV) reporting pipeline, which involves tiering of CNV calls produced by the Canvas software^2^. Although calls produced by the Manta software^3^ are also made available in the GEL research environment, they are not being considered in the GEL’s CNV reporting pipeline.

Here we used SVRare to collate 554,060,126 deletions, duplications and inversions from all the 71,408 participants in the rare disease arm of GEL. From a pilot study of 4313 families, SVRare identified 36 potentially dcSVs; this was equivalent of a 4-fold increase in SV-based diagnostic yield.

We also performed a genome-wide association study, and uncovered clusters of dcSVs in genes with known pathogenicity, such as *PKD1/2* - cystic kidney diseases and *LDLR* - familial hypercholesterolaemia.

All SVs of interest described in this study are detailed in Supplementary table 2. The codebase for SVRare can be found in https://github.com/Oxford-Eye/SVRare

## Method

### Database configuration

SVs had been called using Manta^1^ and Canvas^2^. SV types called by Canvas included LOSS and GAIN, and those called by Manta included DEL, INV (inversion), INS (insertion), DUP (duplication of tandem repeats), and BND (breaking ends). For this work, Canvas LOSS, Canvas GAIN, Manta DEL, Manta INV were included for further analysis, since the similarity calculation is less complex for these types. For the purposes for the current study, Canvas LOSS and Manta DEL are treated as the same type: LOSS.

The workflow is shown in Figure 2. Most samples on GEL were aligned against GRCh38. A small fraction that were aligned against GRCh37, the SV coordinates were first lifted to GRCh38 before being imported to the MySQL database with the other samples. Those that failed the liftover were discarded. Therefore, all the genomic coordinates mentioned in the texts use GRCh38 as the reference, unless otherwise stated. Structural variants were imported into the “Interval” table with the following features: chromosome, start, end and type. Type takes one of the three values: “LOSS” (either called from Manta DEL, or Canvas LOSS), “GAIN” (called from Canvas) and “INV” (called from Manta). Event specific data, such as “sv_id” (the ID value from the Manta or Canvas call), “filter”, source (either Manta or Canvas), genotype (either “HET” or “HOM”), CN (copy number from Canvas call) were imported into the “Participant_Interval”” relation table. Genes that are covered by the SV were recorded in the “Interval_Gene”” relation table. Participant”s HPO terms were imported into the “Participant_HPO” relation table. Details of all the panels in PanelApp, and which panels were applied to which participants, were also imported into the database.

**Figure 2:**
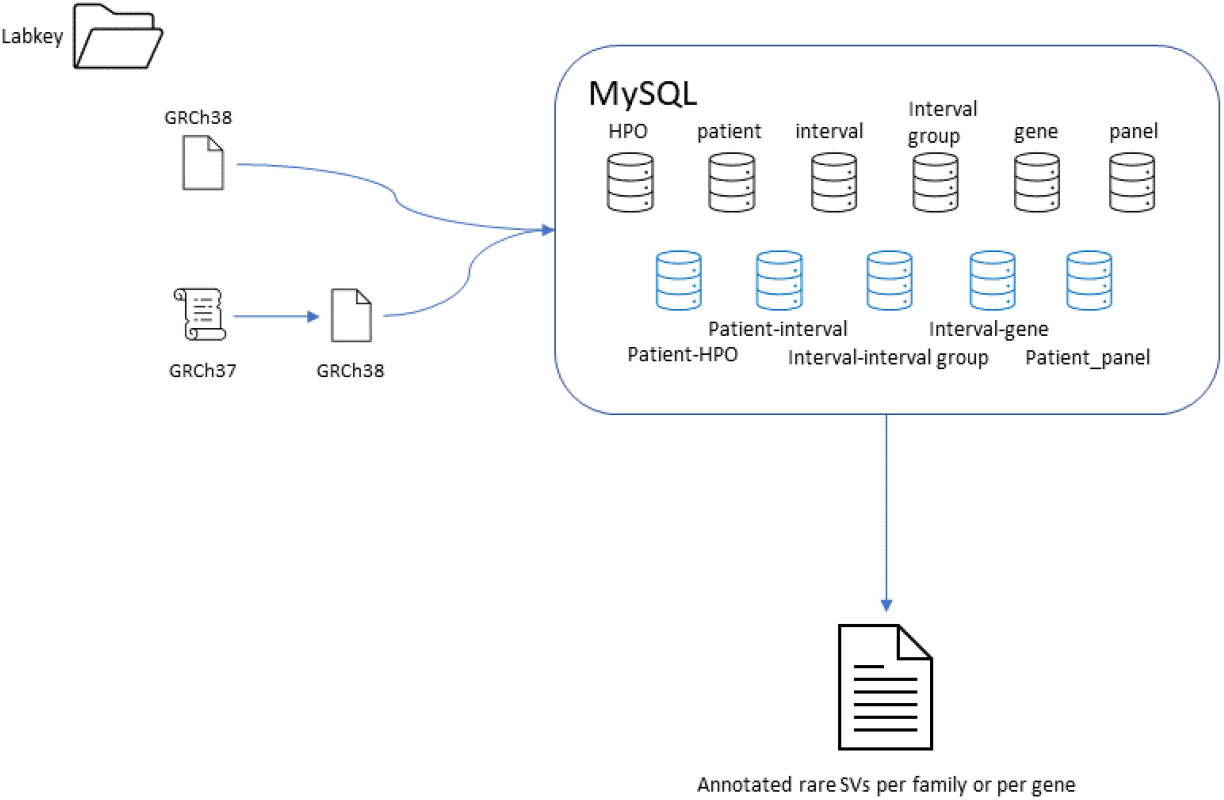
SVRare workflow on GEL. It used Labkey to locate the VCF file for each participant. The VCF file was a merged result from Manta and Canvas calls. If the genome build was GRCh37, it converted the coordinates of all SVs to GRCh38, when possible. The SVs of type LOSS (deletion), GAIN (duplication) and INV (inversion) were imported into a MySQL database, together with participant details, such as disease, HPO terms, family and gene panels that had been applied to the participant. Intervals were then clustered using DBSCAN (eps = 0.1, minimal cluster size = 2). When queried for rare SVs, it would filter for clusters with a small cluster size, then annotate each SV with estimated AF, genes covered, if genes were of interest (either based on HPO terms, or gene panels), if coding regions were disrupted, if the SV was also observed in other family members, and if the SV was also observed in other participants affected by the same disease. Note that some panels in the PanelApps in GEL included genomic regions instead of gene names; they were not imported in the database.

Intervals of the same type were clustered using DBSCAN, with *eps* = 0.1and *min*_*samples* = 2. Similarity between two intervals was calculated as the fraction of the overlap (intersection) over the total length (union). Hence the similarity between any two intervals ranges between 0 and 1. The result of the clustering, together with the sample-size of each cluster, were imported into the tables “Interval_Group” and “Group”, respectively.

### GEL 4K families pilot study

To demonstrate the effectiveness of SVRare in finding disease-causing SVs, we applied it to 4,365 families from the rare disease arm in GEL (both solved and unsolved). Among them, 52 families had missing variant calling files for their proband and were removed from the downstream analyses. This left us with 4313 families. To maximise our chance of identifying pathogenic SVs, we applied the most stringent filters to all the rare SVs found in all the 4313 families: each SV from a family had to satisfy: 1) no other SV with a similarity score of >0.5 can be found carried by an individual who was not a family member and was not affected by the same disease; 2) for deletions, the SV had to cover at least one coding region of a green gene from a gene panel that had been applied to the proband of the family; 3) for inversions and duplications, the SV had to cover at least one coding region, and one of its two ends had to be within a green gene from a gene panel that had been applied to the proband of the family.

### Gene-disease association study

We applied SVRare to all the 19,977 protein coding genes (coordinates extracted from Ensembl Homo_sapien GRCh38 version 98). For each gene, we used DBSCAN to cluster all the SVs, and clusters with a sample size over 50 were filtered. Fisher’s exact test was applied to SVs that are likely to disrupt a coding region of the gene. For this work, only deletions were considered.

## Results

### Summary of SVs in GEL

There were 71,408 participants from 33,923 families in the rare disease arm of GEL (snapshot on the 20th of June, 2021). They included 221 normalised rare disease terms. From them, a total of 554,060,126 SVs were imported to the MySQL database (average 7,759 SVs per individual), among which 415,568,340 SVs had a ‘PASS’ filter inherited from Manta or Canvas (75% PASS rate). Of all the ∼554 million SV records, the majority were recurrences. For example, all of the 71,408 participants carry a chr2:g.186572387_186572581del. There were 20,773,308 distinct SVs, most of which were rare; and only a small fraction of those were carried by >1% of the participants (52,073 SVs, 0.2% of all distinct SVs). Strikingly, these 0.2% common SVs account for 85.5% of all SV calls. Around 72% of all the distinct SVs were deletions (14,853,528), 20% were duplications (4,239,974), and 8% were inversions (1,679,806).

### Pilot study of 4313 families

#### Overview

In order to determine the effectiveness of SVRare, we applied the method to 4313 families (we will refer to it as 4k families henceforth). They encompassed 179 distinct normalised rare disease terms (Supplementary table 1 shows the top 15 diseases). 686 of them had previously been solved or partially solved, of which 9 diagnoses were based on SVs (excluding 2 microsatellite expansions as they are out of the scope of this work); and SVRare correctly prioritised 6 of the 9 SVs. Of all the 3 SVs missed by SVRare, 2 were miss-called by both Manta and Canvas and thus were not present in the SVRare’s database. Both diagnoses were likely made outside of the 100KGP program. The third SV was a deletion in a region with ∼300 other deletions with a similarity score >0.8, and was considered as a ‘common’ SV by SVRare.

A total of 280,285 rare SVs were pooled from the 4k families (mean of 65 SVs per family), of which 35,221 were private (mean of 8 SVs per family). Interestingly, WGS aligned against GRCh37 produced more than double rare SVs than GRCh38, in average (mean: 46 vs 141, median: 35 vs 71, Figure 3A), a likely result of build-specific artifacts. Among all the rare SVs, there were 247,322 deletions (88.2%), 23,822 duplications (8.5%) and 9,141 inversions (3.3%).

**Figure 3:**
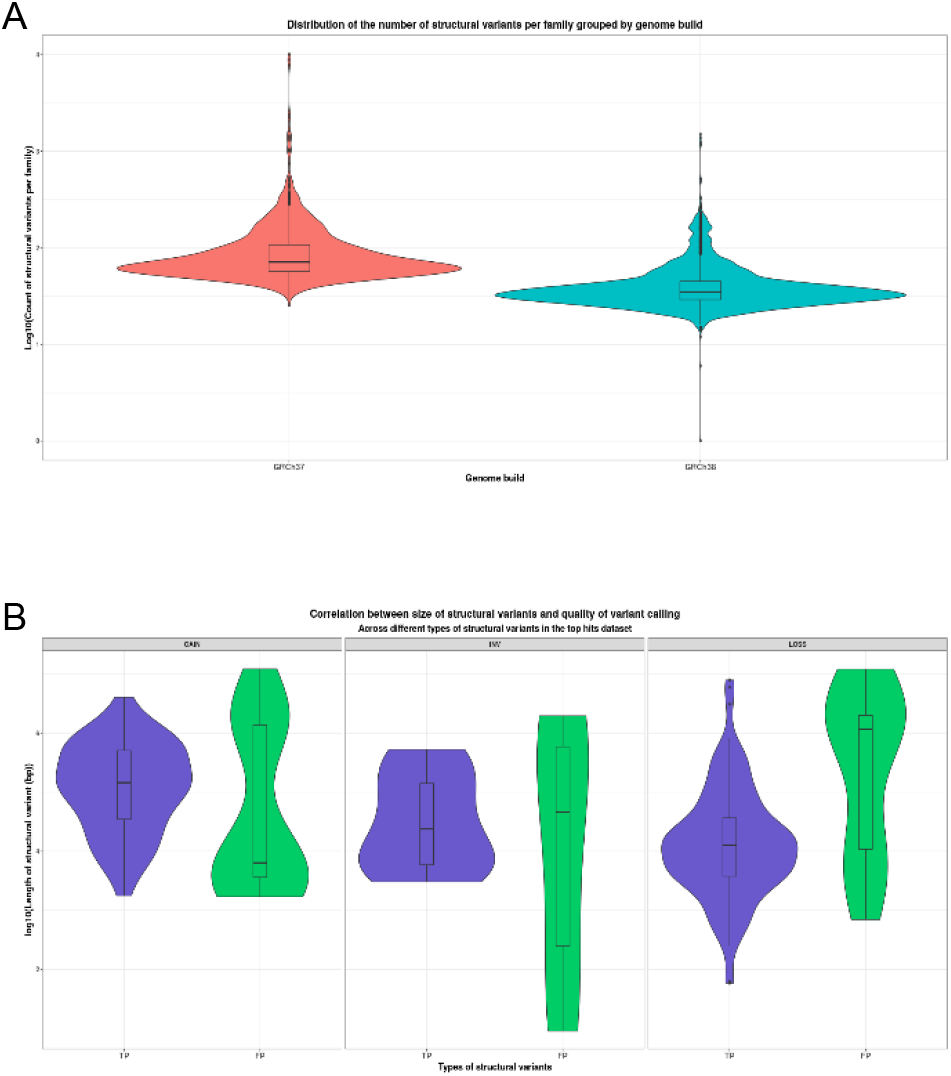
**A)** Distribution of rare SV count per family against genome build from the 4k families. Y-axis is in log10 scale. **B)** Distribution of SV length against true/false positives.SV sizes of false positives are significantly longer than true positives for LOSS/deletions.

In order to find disease-causing SVs, we chose those that were private, disrupt coding regions of a gene, and the gene was a green gene on a panel that had been applied to the proband of a family. This gave 223 SVs in 197 families as a result (one family had 8,832 rare SVs, and exceptionally more private SVs than the average, most of which were false positives. The family was removed from the list). Of them, 132 (59%) were deletions, 74 (33%) were duplications, and 17 (8%) were inversions. We manually inspected each SV and found 176 true positives (precision = 78.9%). As expected from the observation above, precision strongly correlates with genome builds (precision for GRCh38: 87.4%, precision for GRCh37: 59.6%, *p* = 6.8*e*^−6^ of chi-square test of independence). A second variable correlated with precision was the SV size: the larger the size, the higher the likelihood of the SV being false positive, which was significant for deletions (Figure 3B).

#### Complex SVs

Since Manta and Canvas were not able to identify cxSVs, they were usually called as multiple SVs next to each other. From the manual inspection of the 223 SVs, we found three pseudogenes, four inverted deletions (INVdel), one paired-deletion inversion (delINVdel), one paired-deletion translocation (deldelTrans), and one deletion inversion duplication (delINVdup) (Figure 4). The three pseudogenes were unlikely to be disease-causing: they typically appear as complete intronic deletions in the known genes, but they are often inactive elements inserted into regions with unknown clinical relevance.

**Figure 4:**
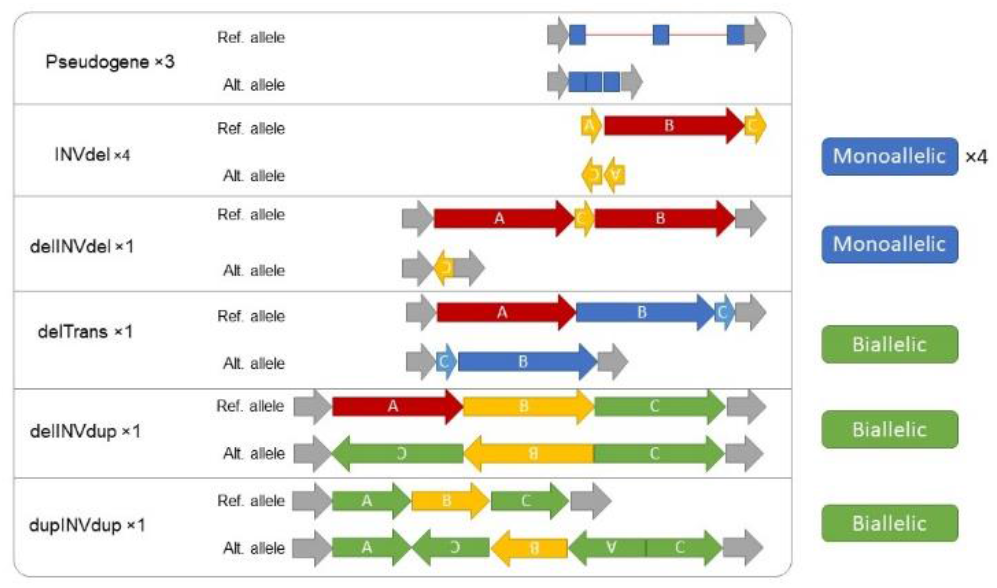
Complex SVs found in the 4k families study. They include pseudogene, inverted deletion (INVdel), paired-deletion inversion (delINVdel), deletion inversion duplication (delINVdup), and paired-duplication inversion (dupINVdup).

The four INVdel were observed in *GRIN2B* – Intellectual disability, *LGI1* – Familial focal epilepsies (Genetic epilepsy syndrome panel), *MEIS2* – Syndromic cleft lip and or cleft palate (Clefting panel, Intellectual disability panel), and *PKD1* – Cystic kidney disease. Interestingly, all four INVdel SVs were associated with autosomal dominant (AD) inheritance. Considering that over all there were 31 rare SVs in genes mostly associated with AD and 87 SVs in genes largely associated autosomal recessive (AR) disorders, this enrichment of INVdel in AD genes associated is very unusual.

The delINVdel was identified in the *DSG2* gene of a participant affected by arrhythmogenic right ventricular cardiomyopathy. Since *DSG2* is associated with autosomal dominant arrhythmogenic cardiomyopathy, this complex SV is likely the cause of the participant”s disease.

The heterozygous delTrans was found in the *NDUFS1* gene of a participant affected by intellectual disability. However, since *NDUFS1* is associated with autosomal recessive intellectual disability, and we were unable to find a second pathogenic variant in the gene, its clinical relevance in this case is therefore uncertain.

Similarly, the delINVdup SV was not considered as causal since the participant carrying the SV in the *UROC1* gene had previously been solved on the *NR2F1* gene for intellectual disability.

A heterozygous paired-duplication inversion was found in the *FAM20C* gene. The 5’ and 3’ end of the inversion were in the 3^rd^ intron of *FAM20C* and downstream of *FAM20C*, thus the inverted copy of *FAM20C* missed the first three exons and was unlikely to encode any protein. However since *FAM20C* is associated with AR disorders, and a second rare variant was not found in this study, the clinical relevance of this cxSV remains unknown (i.e. this individual may be a carrier).

#### Interesting SVs

SVRare made the discovery of pathogenic SVs fast, and we found 97 ‘interesting’ SVs. To be counted as interesting, one SV had to be predicted as protein-coding disrupting, and was only found within the family, or unrelated individuals affected by the same disease. We considered SVs interesting if they were not pseudogenes, the individuals carrying the SVs were stated as unsolved in GMC, and the inheritance of the SVs did not conflict with the family’s pedigree (Table 1). We also merged SVs if they could collectively explain a cxSV event. Notably, four deletions were on genes associated with AD diseases, but were also found to be carried by an unaffected parent (*CYFIP2* – Intellectual disability, *RAC1/ACTB* – Intellectual disability, *ERF* – Craniosynostosis, and *TAOK1* – Intellectual disability). Therefore, the clinical relevance of the SVs was uncertain, and their ‘interesting’ status was assigned with ‘unsure’.

There were 27 interesting SVs predicted to be disrupting protein-coding in genes associated with AD diseases, and an additional 3 chromosome X hemizygous SVs found in XY participants. Among them, 10 were *de novo*, 2 X-linked affected male probands inherited the SVs maternally, and 16 had insufficient information to determine SV hereditary. In the two remaining cases, *TRIO* (Intellectual disability) was inherited from the unaffected mother who was mosaic on the SV, and *UBE3A* (Intellectual disability, a paternally imprinted gene) was maternally inherited.

For the 67 interesting SVs in AR genes, we went to search for a short nucleotide variant (SNV) segregating with the disease. To be considered as pathogenic, the variant must be rare (gnomAD v3.0 allele frequency <= 0.05 or GEL allele frequency <= 0.05, and homozygote/hemizygote count <= 1 in gnomAD) and predicted as likely disease-causing (CADD_phred^4^ >= 15 or MaxEntScan_diff^5^ > 2 or SpliceAI_DS^6^ > 0.5). This resulted in 3 SNVs compound-heterozygous with the SVs of interest. A further 2 SNVs were found in singleton families. It is likely that more biallelic disease-causing changes can be found with further inspections.

A heterozygous SV knocking out exon 23 in *MYO3A* (aa.836-879, NM_017433.5), a gene usually linked to AR hearing loss, was found in three individuals of a family who all had hearing loss (proband, father, and a full sibling). However, recent research illustrated the molecular mechanism underlying an AD form of hearing loss caused by dominant negative mutations in *MYO3A* that encodes the motor domain (aa.338-1053) of the protein^7^. Therefore, we surmise that the heterozygous deletion of exon 23 in *MYO3A* was disease-causing in this AD hearing loss family.

### Gene - disease associations

We grouped rare protein-coding disrupting SVs on a gene level and performed disease associations. For this study, we performed association study on deletions only, as it was recognised as the predominant SV type for causing diseases. As shown in Figure 4, this uncovered many well-established gene-disease relationships, e.g. *PKD1/2* – Cystic kidney disease, *LDLR* – Familial hypercholesterolaemia, and *SPAST* - Hereditary spastic paraplegia. However, majorities of the deletions identified had not been previously reported, and the carriers remained unsolved.

A notable difference between SNP GWAS and SV GWAS is that an SV may span multiple genes, and only one or two of the genes are of clinical relevance. Therefore, it could give rise to false associations on neighbouring genes. A prime example of this was *TSC1* – Classical tuberous sclerosis. Four affected participants from the same family carried the same 1.4Mb deletion that knocked out *TSC1, GFI1B, GTF3C5, CEL, RALGDS*, and *GBGT1*. Although perhaps only *TSC1* was disease-causing, the other neighbouring genes also produced a strong association with Classical tuberous sclerosis.

**Figure 4:**
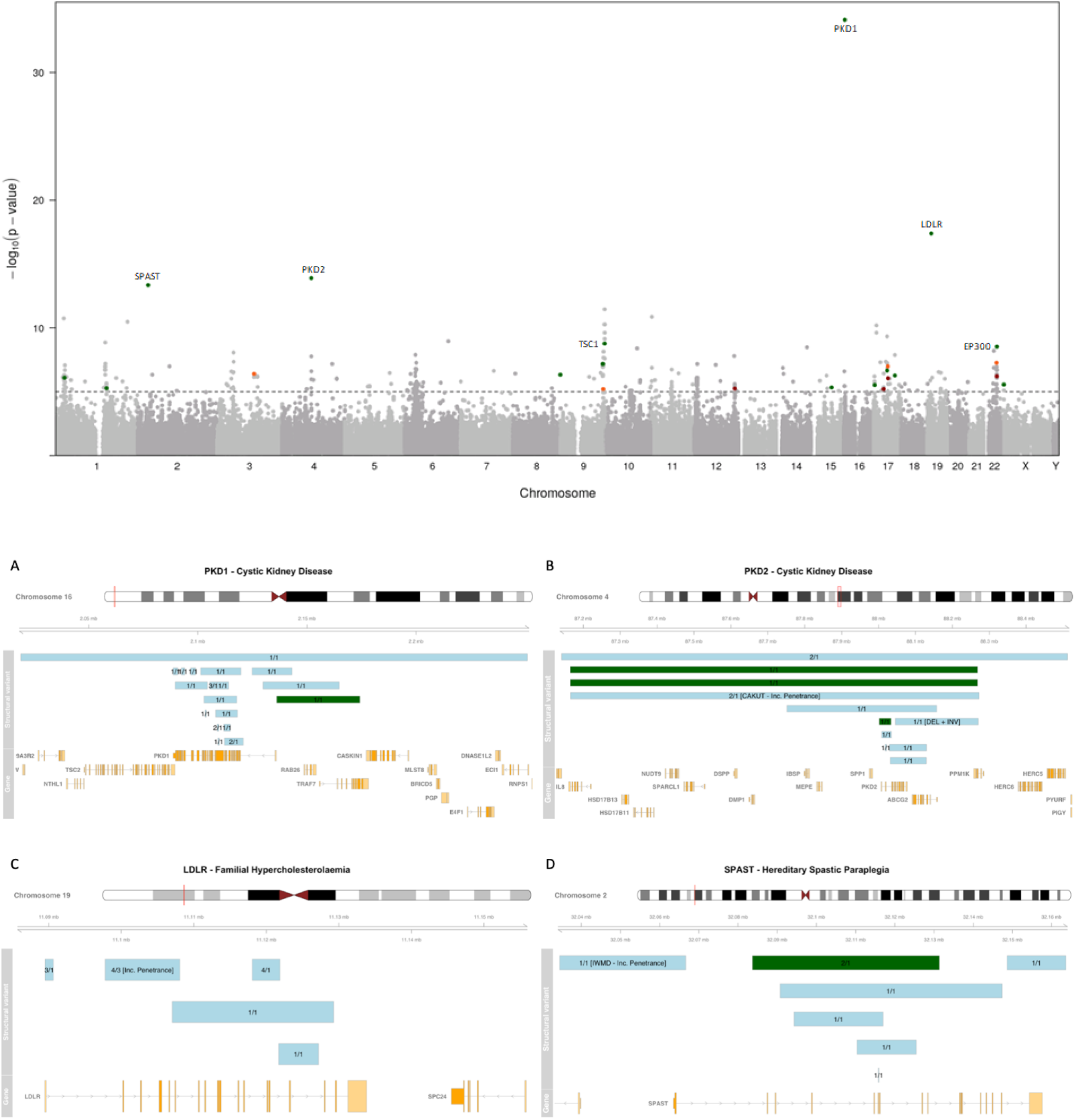
Disease associations. **Top panel:** Manhattan plot shows strong associations in known gene-disease relations. Known gene-disease relations below the significant level of p = e^-5^ are highlighted according to the colour scheme of GEL PanelApp. Annotated are PKD1 (with cystic kidney disease), LDLR (with Familial Hypercholesterolaemia), PKD2 (with cystic kidney disease), SPAST (with Hereditary spastic paraplegia), TSC1 (with classical tuberous sclerosis) and EP300 (with intellectual disability). **Bottom panel**: genome tracks to show SV positions. Green tracks are SVs previously identified. Teal tracks are novel SVs. The numbers on track depict the number of carriers / affected families. A) PKD1 – Cystic kidney disease. B) PKD2 – Cystic kidney disease. C) LDLR – familial hypercholersterolaemia. D) SPAST – Hereditary paraplegia. SV tracks highlighted in green had previously been confirmed as disease-causing. The numbers on each SV track are “number of carriers” / “number of carrier families”. There are two cases with incomplete penetrance and unmatched disease terms: one in B) where the SV is carried by two of a family affected by CAKUT (one is an unaffected relative); the other in C) where the SV was found in three families, and one family was registered with ‘Ultra-rare undescribed monogenic disorders’, and has an unaffected carrier.

We found a 1.2Mb deletion (chr4:g.87164899_88268586del) carried by four participants that knocked out *PKD2*. Two had previously been solved based on the deletion, and the other two were from the same family, with the proband having Congenital Anomaly of the Kidneys and Urinary Tract (CAKUT) instead of Cystic kidney disease. More interestingly, the other carrier from the family was not affected, thus the penetrance of the deletion may not be complete. Although the coordinates were reported differently, a manual inspection of the IGV plots concluded that they were in fact deletions of the identical region. A kinship analysis revealed that the two solved participants were related to the second degree (kinship coefficient of 0.126383)^8^, but neither of them were related to the CAKUT family. Analysis of the surrounding regions reviewed rare SNPs shared only by the four participants, indicating the alleles carrying the deletion share a common ancestor. Further investigation will be required to confirm this hypothesis. Similarly, a 10kb deletion (chr19:g.11097784_11108089del) in *LDLR* that knocks out most of its ligand binding domain^9^ was carried by four participants. Two unrelated individuals were affected by typical Familial hypercholesterolaemia; but the other two were from a family that had a normalised disease term of “Ultra-rare undescribed monogenic disorders”, with one of them being unaffected. These two cases demonstrated that same SVs may confer varying phenotypes.

### Three solved cases

During the course of this study, three cases recruited in Oxford were solved based on the results produced by SVRare.

### *REEP6, CNGB3* and retinal dystrophy

As a result of initial testing of SVRare in a group of participants recruited in the Oxford Eye Hospital, we identified a homozygous deletion in the *REEP6* gene, and a heterozygous tandem duplication in the *CNGB3* gene. The *REEP6* deletion (chr19:g.1490941_1492690del) knocks out the first exon of the gene, and the *CNGB3* SV (chr8:g.86652138_86663932dup) duplicates exon 7. We also identified a second variant in *CNGB3* (chr8:g.86643780AG>A, NP_061971.3 p.Thr383IlefsTer13). Given the phenotype is supportive in both cases (Figure 5), we surmise that the SVs are highly likely to be disease-causing.

**Figure 5:**
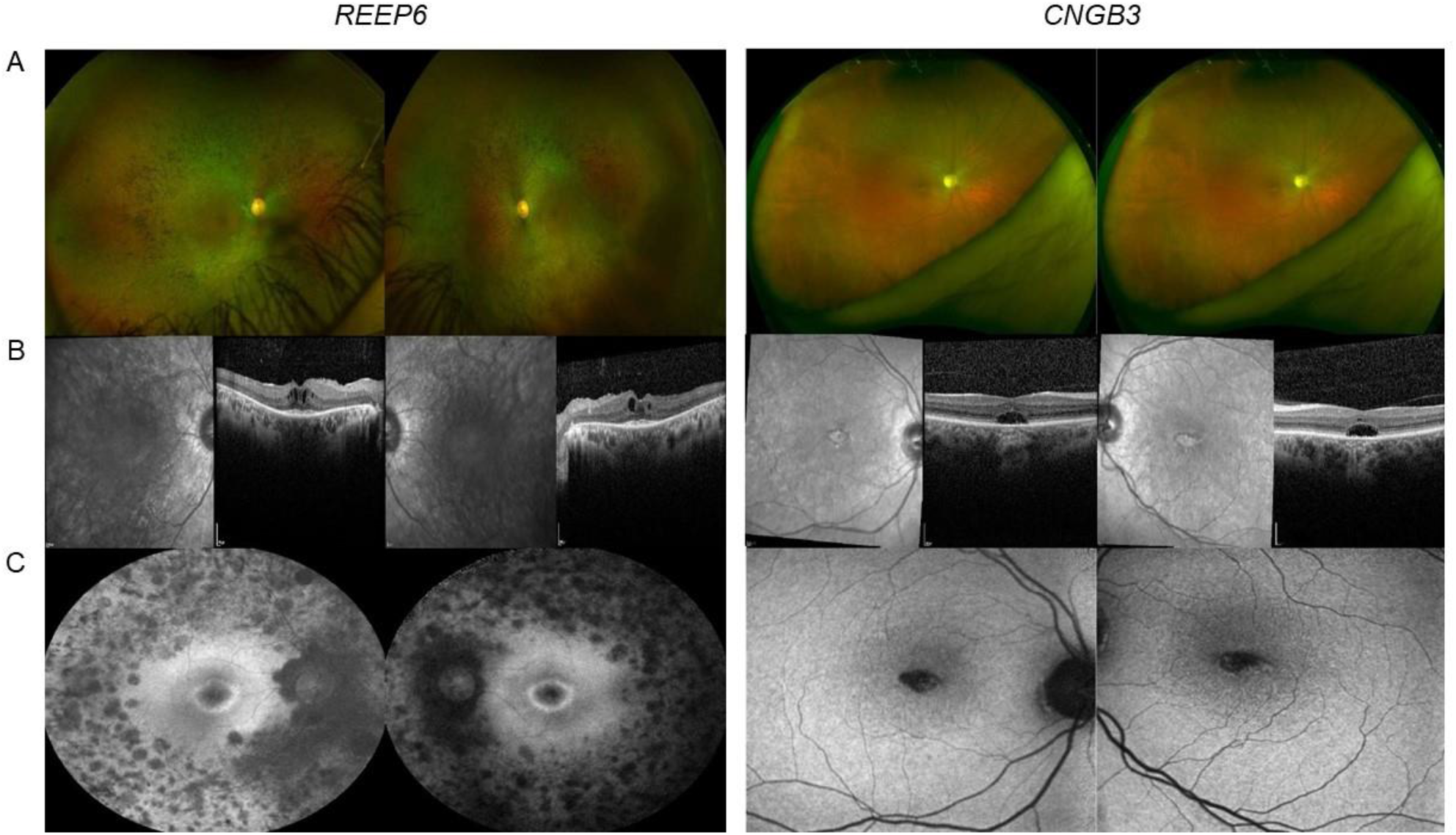
retinal imaging of patients carrying a homozygous REEP6 deletion and a heterozygous CNGB3 tandem duplication. Panel **A** shows colour imaging of the REEP6 patient that displays bilateral peripheral bone-spicule pigmentation mainly located in the mid periphery, with attenuated blood vessel and pale optic discs; and of the CNGB3 patient that displays a central patch of atrophy involving both central maculae, with mild pallor of discs and subtle attenuation of vasculature. Panel **B** shows spectral domain optical coherence tomography (SD-OCT) of the REEP6 patient that displays bilateral cystoid macular oedema, atrophy of the outer retinal layers external to the fovea with thickening of the ellipsoid zone at the fovea; and of the CNGB3 patient that displays bilateral optical gaps with loss of the ellipsoid zone involving both foveae. Panel **C** shows short-wavelength autofluorescence imaging of the REEP6 patient that displays an annulus of increased signal around the fovea with widespread patchy loss of signal corresponding to areas of atrophy in the mid periphery; and of the CNGB3 patient that displays central gross reduction in signal involving both central maculae consistent with atrophy with an irregular edge and a small patch of retained AF signal at both foveae.

### *KDELR2* and Osteogenesis imperfecta

A family-oriented analysis was performed on a parent-proband-sibling family from 100KGP where both the proband and her brother presented with severe Osteogenesis Imperfecta (OI).

In the proband, we identified two compound heterozygous intragenic deletions in *KDELR2* (Supplementary Figure 6). The affected brother also harboured the same combination of deletions, with phasing confirmed by parental inheritance. This gene was only associated with OI in 2020, when four families with biallelic SNVs were identified^11^. A more recent follow up study identified two further families with OI and additional neurodevelopmental features^12^. This is also the first reported OI case that was caused by biallelic SVs. Detailed clinical and neurodevelopmental assessment of the family described here is underway and will be described elsewhere.

**Figure 6:**
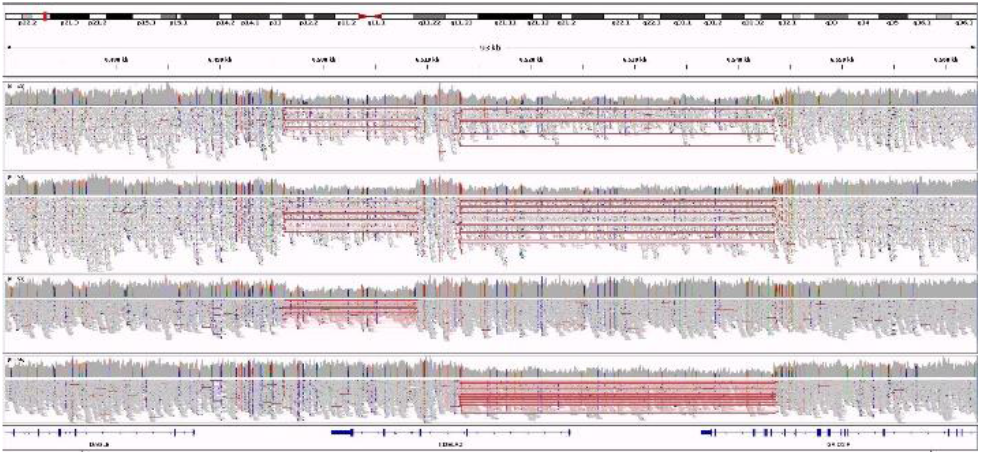
An Integrative Genomics Viewer shows two exonic deletions disrupting KDELR2 in a family affected by Osteogenesis imperfecta. Top two tracks were affected siblings. Bottom two tracks were unaffected parents.

## Discussion

Many tools have been developed to call SVs from short-read WGS^10^. However, high false-positive rates and a lack of means to efficiently merge SVs from different individuals have been hindering dcSV discovery. Indeed, from the 686 existing diagnoses or partial diagnoses in the 4k families included in our pilot study (15.9%), SV contributed to only nine, four of which were likely made outside GEL’s diagnostic pipeline.

In this study, we introduce SVRare and its implementation in 100KGP that enabled fast dcSV discovery. It aggregated 554 million SV events called from Manta and Canvas in 71 thousand individuals, or 34 thousand families. In the 4k families pilot study, we used SVRare to identify 97 interesting SVs, with 31 of them likely causing AD (or X-linked) disorders. We also found a second rare variant predicted to be pathogenic in five AR cases that segregates with an interesting SV. Suppose all the 36 SVs are causal, this is a four-fold increase in SV-based diagnostic yield. From the 9 SV-based diagnoses previously made, SVRare identified 6 of them. Excluding two that were miss called by Canvas and Manta, SVRare achieved a high recall (85.7%).

Notably, all the 9 dcSVs identified prior to SVRare were deletions. In contrast, SVRare uncovered a range of different types of dcSVs in the pilot study, including 64 deletions, 23 duplications, three inversions, and seven cxSVs. In the 31 SVs associated with AD disorders, 18 were deletions, 6 were duplications, 2 were inversions and 5 were cxSVs. Interestingly, all the 5 cxSVs involving both inversions and deletions found in this study were associated with AD disorders. The reason for the unbalanced distribution of cxSVs underlying AD and AR disorders remains unknown. Further studies will be carried out to verify this finding.

The 4k family pilot study was conducted to consider only private protein-coding disrupting SVs: an SVs was considered private if they were only found in families that had the same normalised disease term. This setting inevitably favoured the findings associated with AD disorders, since protein-coding disrupting SVs in genes associated with AR disorders tend to be more tolerated, and thus are more pervasive in the general population. It is therefore highly likely that a dedicated family-wise investigation will produce more findings.

In this study, we performed GWAS for deletions only, since it was the predominant SV type in the database. As shown in Figure 4, many of the top ranked associations were well known, such as *PKD1/2* and Cystic kidney disease. However, majority of the deletions underlying the strong associations had not been reported.

An advantage of gene-wise analysis over family-wise analysis is that one can systematically investigate all dcSVs within a gene. For example, the heterozygous deletion that completely knocked out *PKD2* was not reported in a carrier with CAKUT. We suspect that this was because the unaffected relative of the proband was also a carrier of the deletion. From our gene-wise analysis on the gene, we found that the same deletion had been confirmed disease-causing in two separated cases. We surmise that the whole gene deletion of *PKD2* has incomplete penetrance. We also uncovered a list of rare SNVs shared exclusively among the four carriers, indicating a common haplotype carrying the deletion. The rare SNVs can then be used as a surrogate to probe for the existence of the deletion.

The first step of common practices when investigating SVs is usually removing calls that do not pass the filter in variant calling tools such as Manta and Canvas. However, in the 4k families pilot study, we found that 31 out of the 97 interesting SVs failed either Manta or Canvas filters. Therefore, we suggest against removing SVs based on filters. Further investigation is required to study the distribution of different filters in the true and false positives.

One problem with merging records aligned against GRCh37 and GRCh38 was that a proportion of SVs were dropped during the process (i.e. LiftOver tools fails for regions that do not map 1 to 1). Despite this, samples aligned against GRCh37 produced significantly more rare SVs in average (Figure 3A). The reason for this is still unclear, but we suspect that it is due to genome-build related artifacts. The solution would be to realign all GRCh37 samples to GRCh38 and recall SVs.

## Data Availability

All data produced in the present study are available in the manuscript and online at https://github.com/Oxford-Eye/SVRare

## Acknowledgement

We would like to thank RetinaUK for supporting fundings for JY’s work on SVRare; John Fell Fund for setting up a local GEL user group which made this work possible. This research was also supported by the National Institute for Health Research (NIHR) Oxford Biomedical Research Centre Programme, and the Wellcome Trust (203141/Z/16/Z). This research was made possible through access to the data and findings generated by the 100,000 Genomes Project. The 100,000 Genomes Project is managed by Genomics England Limited (a wholly owned company of the Department of Health and Social Care). The 100,000 Genomes Project is funded by the National Institute for Health Research and NHS England. The Wellcome Trust, Cancer Research UK and the Medical Research Council have also funded research infrastructure. The 100,000 Genomes Project uses data provided by patients and collected by the National Health Service as part of their care and support

## Supplemental materials

**Supplementary table 1.** Disease count of the top 15 diseases in the pilot 4k families

| disease | count |
| --- | --- |
| Intellectual disability | 1021 |
| Primary immunodeficiency | 230 |
| Cystic kidney disease | 168 |
| Ultra-rare undescribed monogenic disorders | 145 |
| Familial Thoracic Aortic Aneurysm Disease | 137 |
| Proteinuric renal disease | 90 |
| Familial breast and or ovarian cancer | 85 |
| Familial Hypercholesterolaemia | 81 |
| Congenital hearing impairment | 79 |
| Hereditary ataxia | 74 |
| Epileptic encephalopathy | 70 |
| Hypertrophic Cardiomyopathy | 65 |
| Multiple Tumours | 58 |
| Congenital Anomaly of the Kidneys and Urinary Tract (CAKUT) | 53 |
| Hereditary Spastic Paraplegia | 51 |

**Supplementary table 2**, SVs of interest discovered in the pilot 4k families

https://github.com/Oxford-Eye/SVRare/raw/master/supplementaries/top_hits_for_publish.xlsx

